# Oxygen-based autoregulation indices associated with clinical outcomes and spreading depolarization in aSAH

**DOI:** 10.1101/2024.05.17.24307563

**Authors:** Andrew P Carlson, Thomas Jones, Yiliang Zhu, Masoom Desai, Ali Alsarah, C. William Shuttleworth

## Abstract

**Background:** Impairment in cerebral autoregulation has been proposed as a potentially targetable factor in patients with aneurysmal subarachnoid hemorrhage (aSAH), however there are different continuous measures that can be used to calculate the state of autoregulation. In addition, it has previously been proposed that there may be an association of impaired autoregulation with the occurrence of spreading depolarization (SD) events.

**Methods:** Subjects with invasive multimodal monitoring and aSAH were enrolled in an observational study. Autoregulation indices were prospectively calculated from this database as a 10 second moving correlation coefficient between various cerebral blood flow (CBF) surrogates and mean arterial pressure (MAP). In subjects with subdural ECoG (electrocorticography) monitoring, SD was also scored. Associations between clinical outcomes using the mRS (modified Rankin Scale) and occurrence of either isolated or clustered SD was assessed.

**Results:** 320 subjects were included, 47 of whom also had ECoG SD monitoring. As expected, baseline severity factors such as mFS and WFNS (World Federation of Neurosurgical Societies scale) were strongly associated with the clinical outcome. SD probability was related to blood pressure in a triphasic pattern with a linear increase in probability below MAP of ∼100mmHg.

Autoregulation indices were available for intracranial pressure (ICP) measurements (PRx), PbtO2 from Licox (ORx), perfusion from the Bowman perfusion probe (CBFRx), and cerebral oxygen saturation measured by near infrared spectroscopy (OSRx). Only worse ORx and OSRx were associated with worse clinical outcomes. ORx and OSRx also were found to both increase in the hour prior to SD for both sporadic and clustered SD.

**Conclusions:** Impairment in autoregulation in aSAH is associated with worse clinical outcomes and occurrence of SD when using ORx and OSRx. Impaired autoregulation precedes SD occurrence. Targeting the optimal MAP or cerebral perfusion pressure in patients with aSAH should use ORx and/or OSRx as the input function rather than intracranial pressure.

## Introduction

Management of delayed cerebral ischemia^1^ (DCI) after aneurysmal subarachnoid hemorrhage (aSAH) remains both a significant challenge and one of the major targetable secondary injury mechanisms in the neuro-intensive care unit. Evolving understanding of the mechanisms of DCI have opened new physiologically based therapeutic approaches to prevent and treat this problem. Two of the most relevant of these include the role of impaired autoregulation^2–9^ and occurrence of spreading depolarization (SD) events^7,10–13^.

Cerebral autoregulation (CA) is the adaptive mechanism by which the brain maintains constant cerebral blood flow (CBF) over a wide range of mean arterial pressure (MAP)^14^. In cases of injury, autoregulation can be impaired, shifted, or even lost such that moderately low blood pressure could lead to potentially ischemic levels of blood flow and moderate elevations could lead to hyperemia, elevated intracranial pressure, and secondary damage^3,15^. There is no perfect tool to measure autoregulation, especially in a condition such as aSAH where there can be significant temporal and regional changes in autoregulation the weeks following the initial bleed^16^. Continuous indices have been developed which use a rolling correlation coefficient between MAP and various surrogates for CBF to gain insight into the autoregulatory status and how it evolves over the course of admission^16–18^. A target that minimizes the index can then be hypothesized on a patient-specific basis and potentially be used to refine the patient-specific optimum MAP (MAPopt) or optimum cerebral perfusion pressure (CPPopt)^19^.

SD are massive, non-synaptically mediated, slow-moving depolarizing events that are electrophysiologically very similar to terminal depolarization/ brain death, so can be considered a type of “near death event”^20–22^ especially when occurring in metabolically compromised regions. When there is adequate metabolic substrate, (blood flow, glucose, oxygen) tissue can slowly recover over minutes to hours, beginning neuronal transmission after a transient period of minutes^23^. However, in extremely metabolically compromised tissue, SD can result in expansion of ischemia due to the large metabolic requirements for recovery^22,23^. This process is likely cyclical, where, in vulnerable tissue, metabolic transients such as hypoxia, hypotension, and even cortical excitation can trigger SD^24–26^ and SD can in turn, further stress this metabolically compromised tissue, resulting in expansion of ischemia^27^. In aSAH, the occurrence of SD has been strongly associated with worse clinical outcomes and episodes of neurologic deficit^13^.

An important factor contributing to initiation of SD is inadequate CBF^28,29^. In conditions of impaired autoregulation, where CBF may fall into ischemic zones and trigger SD, even in the normal MAP range, it would be expected that SD may occur more frequently^7^. Several previous studies demonstrated a triphasic probability curve of SD versus MAP where the probability of SD increased dramatically at the lower end of blood pressure, was flat at the middle range, and decreased further at the high end of blood pressure^7,26^. This relationship follows the characteristics of the autoregulatory curve^16^.

We previously assessed a small group of patients with aSAH who all had simultaneous monitoring of multiple different autoregulation indices to assess both agreement between those indices and relative predictive value for clinical outcomes and occurrence of SD^7^. In that study, we found that PRx (derived from ICP), ORx (derived from PbtO2), and OSRx (derived from scalp NIRS) seemed to have the most consistent association with SD and possibly with clinical outcomes, though the study lacked power. In this current study, we expanded this cohort to a much larger data set and performed a more rigorous high-resolution assessment of each autoregulation index to determine whether these indices were associated with worse clinical outcomes and if SD occurrence was the cause or the result of impaired autoregulation.

## Methods

### Subjects

Subjects enrolled in multiple studies related to multimodality monitoring at our institution over a period of 12 years were pooled for the current study (UNM IRB# 10-159, 17-297, 20-390, and 21-044) and data on multimodality monitoring were collected on these patients. All of these were observational studies without research interventions related to SD or multimodal monitoring parameters. Subjects with aSAH were included who had placement of multimodality monitoring. The clinical criteria for placement of multimodal monitoring were any patient with symptomatic hydrocephalus on admission or need for CSF (cerebrospinal fluid) diversion during a surgical procedure. The Hummingbird (IRRAS USA: San Diego, CA) system^30^ was used in this entire cohort. This system consists of a single twist drill bolt with an external ventricular drain (EVD) with an integrated parenchymal monitor for continuous intracranial pressure (ICP) measurement built into the catheter. There are one or two additional side ports that allowed for the placement of additional monitors, typically a PbtO_2_ monitor (Licox: Integra LifeSciences: Princeton, NJ) and a CBF monitor (Bowman Perfusion Monitor: Hemedex: Waltham MA). Most monitors are placed contralateral to the site of aneurysm rupture in case surgical treatment was needed. All patients also had bilateral frontal NIRS (INVOS: Medtronic: Minneapolis, MN) placed for clinical management. Some patients undergoing surgical treatment had additional placement of a 1x6 subdural strip electrode over the region deemed to be at the highest risk of ischemia (e.g. temporal for middle cerebral artery aneurysms or frontal for anterior communicating aneurysms). All these parameters, in addition to systemic parameters such as blood pressure, are monitored at the bedside in the Moberg CNS monitor (Natus: Middleton, WI).

### Scoring

In our previous study, we retrospectively calculated low-resolution versions of the various autoregulation indices using one-minute binned data exported from the Moberg CNS as a 30-minute rolling average of the Pearson’s correlation coefficients between MAP and each potential CBF surrogate^7^. With recent upgrades to the CNS Envision software (Natus: Middleton, WI), the originally proposed method for calculating PRx^31^ can be applied to each additional parameter using the original waveform data. Briefly, each index is calculated as the 10s rolling average of the Pearson’s correlation coefficients between the waveform level MAP and the input function of interest over a 5-minute moving window. We used ICP from the parenchymal monitor to calculate PRx-parenchymal, ICP from the EVD to calculate PRx-EVD, CBF from the Bowman probe to calculate CBFRx, PbtO_2_ from the Licox to calculate ORx, and cerebral regional hemoglobin oxygen saturation (rSO2) from the INVOS to calculate OSRx. We calculated each of these continuous indices using the CNS Envision software for all the retrospective stored subject data.

SD recordings were acquired and scored per international consensus guidelines^20^. Briefly, a standard platinum 1X6 electrode was placed in the subdural space at the time of surgery. ECoG was recorded using a DC amplifier (Moberg CNS) directly into the Moberg CNS system, linking these data to the other multimodal monitoring parameters^32^. DC recordings (high-pass filtered at 0.005Hz to stabilize drift) were overlayed on high-frequency bandpass filtered data (0.5-50Hz). A SD cluster includes all SDs in the same patient occurring within 1 hour of a consecutive pair.

### Data processing

The steps for data processing to a workable format required extensive development and are summarized in Figure 1. Briefly, clinically recorded files were first copied from a clinical server to a research folder per IRB requirements. After calculation of autoregulation indices, data were then exported in text files in one-minute bins. Annotations including SD scoring and other events were exported as a separate file. Naming for variables was then standardized from various conventions that changed over the years. In cases where the label was changed during the recording (e.g. ART changed to ABP), these were manually reviewed and then consolidated if it was determined to be the same data stream in that subject. Results for complex cases were manually reviewed to ensure accuracy. In some subjects, there were multiple files which were re-linked into one contiguous file. The recordings were then linked to the date and the times of SD. Data were then filtered to include only physiologically plausible ranges (e.g. to remove times when arterial lines were being accessed or “zeroed”.)

**Figure 1:**
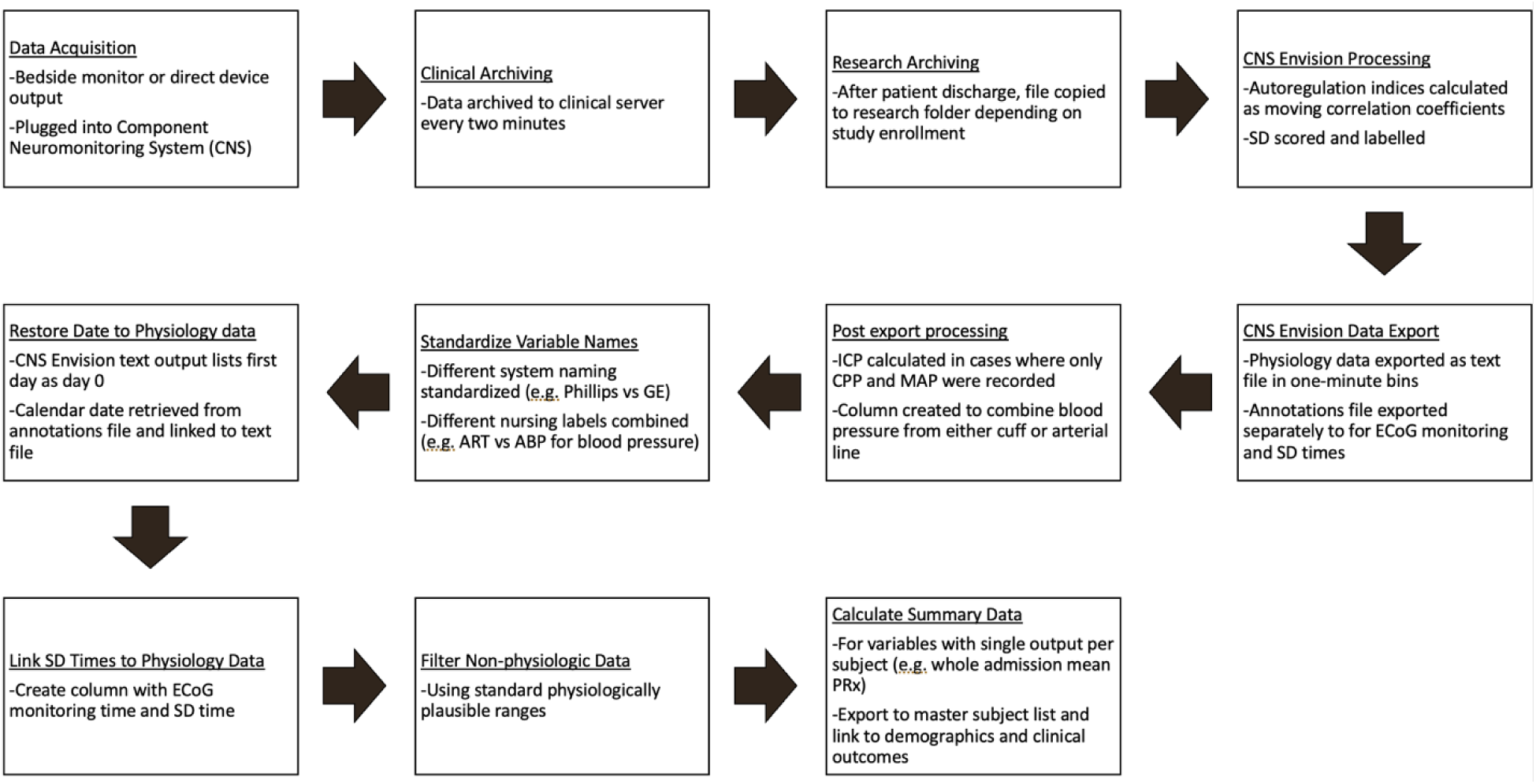
Workflow for data cleaning and analysis.

### Clinical data

Standard clinical variables were recorded either retrospectively with chart review or prospectively collected. Age, sex, admission diagnosis, admission Glasgow coma scale (GCS) score, admission world federation of neurosurgical societies grade (WFNS), and modified Fisher scale (mFS) were all collected. Angiographic vasospasm was recorded as the maximum severity recorded on the routine day 7 angiogram. Discharge and ∼day 90 modified Rankin scale (mRS) were collected either from chart review or from prospective structured interview.

### Data analysis

Summary values for each parameter of interest in addition to clinical and demographic and outcome data were then compiled. The primary clinical outcome was discharge mRS as this was the outcome parameter with the least missing data. Good outcome was defined as mRS 0-3 whereas poor outcome was defined as mRS 4-6. Two-tailed t-tests as well as Mann-Whitney non-parametric tests were used for continuous variables, and Fisher’s test were used for binary variables. Subjects with missing data for a given variable were excluded from that analysis. Autoregulation indices were derived on the basis of Pearson’s correlation and hence are bounded between −1 and 1. We applied Fisher’s transformation for correlation coefficients to the autoregulation indices before conducting t-tests for the difference between poor and good outcomes and pre-SD and post-SD trend analysis. Significance was considered at p-values <0.05.

In order to assess the relationship between SD and blood pressure, probability curves were constructed using 20-minute bins containing SD compared to bins without SD in reference to MAP. To determine the temporal relationship between SD and various autoregulation indices, we investigated the time-trend of these indices pre-SD and post-SD. We first defined and identified SD clusters within a patient using the working definition described earlier. SDs that were more than one hour before the first SD or one hour later after the last SD in a cluster were defined as belonging to different clusters. For each SD cluster, the pre-SD time series begins at 60 minutes before and ends at the first SD of the cluster, with a total of 60 bins including the one for the first SD. The post-SD time series begins at the last SD of the cluster and ends 60 minutes later. These bins were non-overlapping. We applied Fisher’s transformation of correlation coefficients and then conducted trend analysis on the transformed data using linear mixed-effects model with random effects for individual patients as well as SD clusters. We conducted separate trend analysis for non-clusters of single SDs, clusters of multiple SDs, and the two pooled. We also explored other time boundaries such as 2 hour and 30 minutes as well as varying length of the serial data. All processing and analysis were performed using Matlab, R, and Graphpad Prism(v10.1.1). The STROBE reporting guideline tool was used for this observational study.

## Results

We identified 320 subjects meeting the inclusion criteria between 2010 and 2023. Mean Age was 57 (StDev=14). The median admission GCS score was 12 and WFNS score was 3. The overall mean hospital length of stay was 24 days, consistent with these being a relatively poor grade group of patients with aSAH. More patients (n=198) underwent craniotomy or craniectomy and the remainder (n=122) underwent endovascular embolization. This trend has been changing with time as endovascular therapy has improved, however, surgical treatment was preferred especially early in the experience. Forty-seven of these subjects underwent SD monitoring.

Older age, lower admission GCS, higher admission WFNS, and higher admission mFS were all associated with worse outcomes. Sex was not. Interestingly, angiographic vasospasm severity on routine day 7-11 angiogram was also not associated with outcomes. See Table 1.

**Table 1.** Associations of Patient Characteristics, Injury Severities, and Autoregulation Indices with Clinical Outcome at Discharge.

|  | **Good outcome at DC**<br>**(mRS 0-3) N=118** | **Poor outcome at DC**<br>**(mRS 4-6) N=202** | **p-value** | **test** |
| --- | --- | --- | --- | --- |
| **Age in years: Mean (StD)** | 51.81 (12.26) | 60.46 (13.46) | **<0.0001** | t-test |
| **Female Sex: N (%)** | 70 (59.3%) | 139 (68.8%) | 0.0898 | Fisher's exact test |
| **Admission GCS: Median** | 15 (n=29) | 10 (n=94) | **<0.0001** | Mann Whitney test |
| **Admission WFNS: Median** | 1 (n=28) | 4 (n=87) | **<0.0001** | Mann Whitney test |
| **Admission mFS: Median** | 4 (n=24) | 4 (n=58) | **<0.0001** | Mann Whitney test |
| **Vasospasm (score=0-3\*): Median** | 0 (n=103) | 0 (n=157) | 0.2991 | Fisher's exact test |
| **PRx EVD:**<br>**Mean (StD)** | 0.11 (0.10) (n=68) | 0.09 (0.13)(n=122) | 0.2546 | t-test** |
| **PRx parenchymal:**<br>**Mean(StD)** | 0.17 (0.12) (n=68) | 0.18(0.14)(n=132) | 0.6869 | t-test** |
| **ORx:**<br>**Mean (StD)** | 0.03 (n=95) | 0.06 (n=173) | **0.0005** | t-test** |
| **OSRx (average R/L):**<br>**Mean (StD)** | 0.05 (n=98) | 0.12 (n=167) | **<0.0001** | t-test** |
| **CBFRx:**<br>**Mean (StD)** | 0.25 (n=66) | 0.25 (n=103) | 0.9551 | t-test** |

\* 0= no vasospasm, 3=severe vasospasm
\*\*For each autoregulation index Fisher's transformation of correlation coefficient was applied to the data before two-sample t-test.
DC= Discharge, mRS= modified Rankin Scale, StD= Standard deviation, GCS=Glasgow Coma Scale score, WFNS= World Federation of Neurosurgical Societies Grade, mFS= modified Fisher Scale, PRx= Pressure reactivity, EVD= external ventricular drain, ORx= Oxygen reactivity, OSRx= Oxygen saturation reactivity, CBFRx= Cerebral blood flow reactivity

We hypothesized that impaired autoregulation (higher autoregulation index) would be associated with poor outcomes but found only ORx (p=0.0005) and OSRx average (p<0.0001) demonstrated a significant association with worse clinical outcomes (see Table 1 and Figure 2).

**Figure 2:**
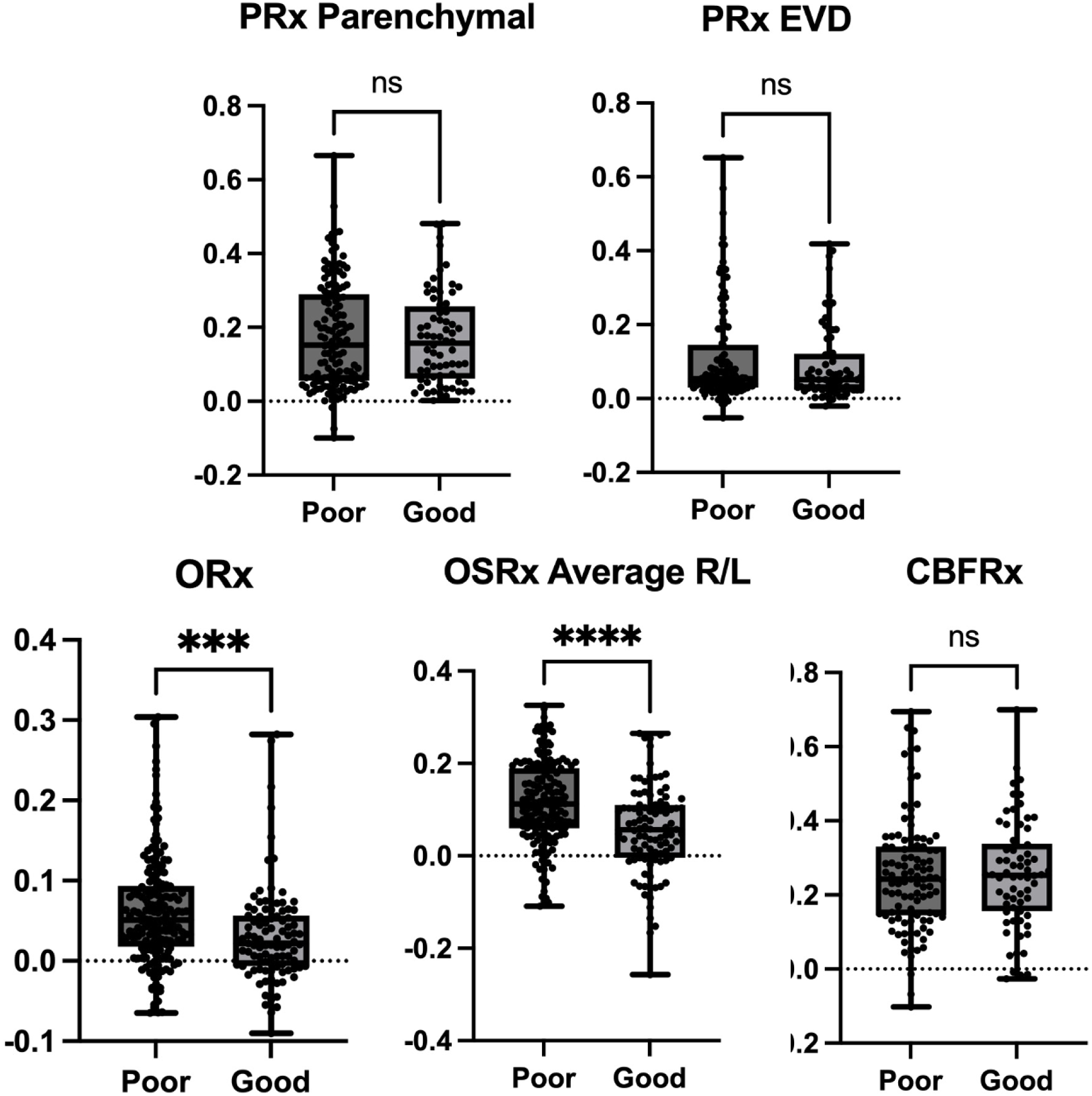
Box plots with raw data for each autoregulation index and clinical outcomes.

Plotting the probability of SD versus blood pressure, a familiar triphasic curve characteristic of cerebral autoregulation was demonstrated^16^ (Figure 3). Thus, below MAP of ∼90mmHg there was a nearly linear association of increased SD probability with decreased MAP. As noted in the plot, between 90 and 150mmHg, the probability of SD was relatively stable and low. Above MAP of 150 no SD were observed. These three transitions have previously proposed to represent a reflection of the autoregulatory curves and the upper and lower limits of autoregulation. In this cohort of poor grade patients overall, a shift in the lower limit of autoregulation may be suggested.

**Figure 3:**
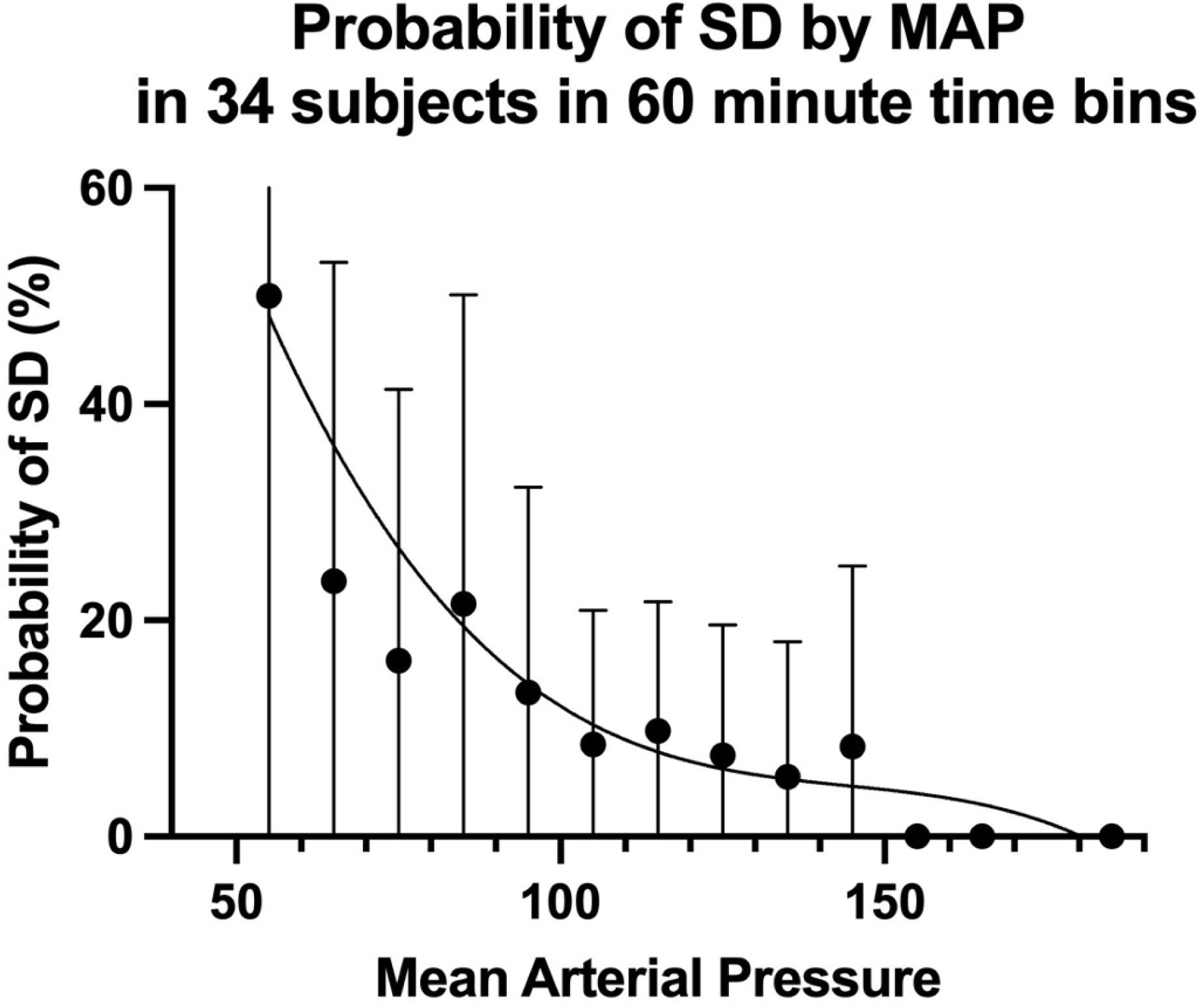
Relationship between probability of SD and MAP. This triphasic curve could plausibly be a reflection of a shifted autoregulation curve in this cohort.

With regard to SD clusters, we found an increasing trend in ORx and OSRx average within 60-min just before SD clusters as seen in the positive slopes derived from the fitted linear mixed-effects model (0.0013, p=0.001 for ORx, and 0.0009, p=0.0001 for OSRx, respectively, Table 2) We also found a decreasing trend in ORx within 60 minutes just after SD clusters (−0.0008, p=0.0288), but no significant trend in OSRx post-SD clusters. Comparable and consistent results were seen when we analyzed single SD clusters and multiple SD clusters separately (See slopes in Table 2). Figure 4a and Figure 4b display this trend. This may suggest that worsening autoregulation measured by ORx and OSRx contributed to increased SD probability with potential improvement (e.g. in ORx) or stabilization (e.g. in OSRx) post-SD. Mean PRx from the parenchymal monitor was significantly higher prior to clustered SDs compared to isolated SD, but with a somewhat decreasing trend immediately before clustered SDs (slope=-0.0015, p<0.0001) and increasing trend post isolated SDs (slope=0.0008, p=0.002). We also found a decreasing trend in PRx EVD prior to SD, but an increasing trend post-SD with only isolated SD. Significant pre-SD and post-SD trends were found in CBFRx only with clustered SDs not isolated SDs. It is interesting to note that the time trends in these autoregulation indices were robust with respect to time boundary, length of index serials, or removing time bins immediately proximate to SD clusters.

**Figure 4.**
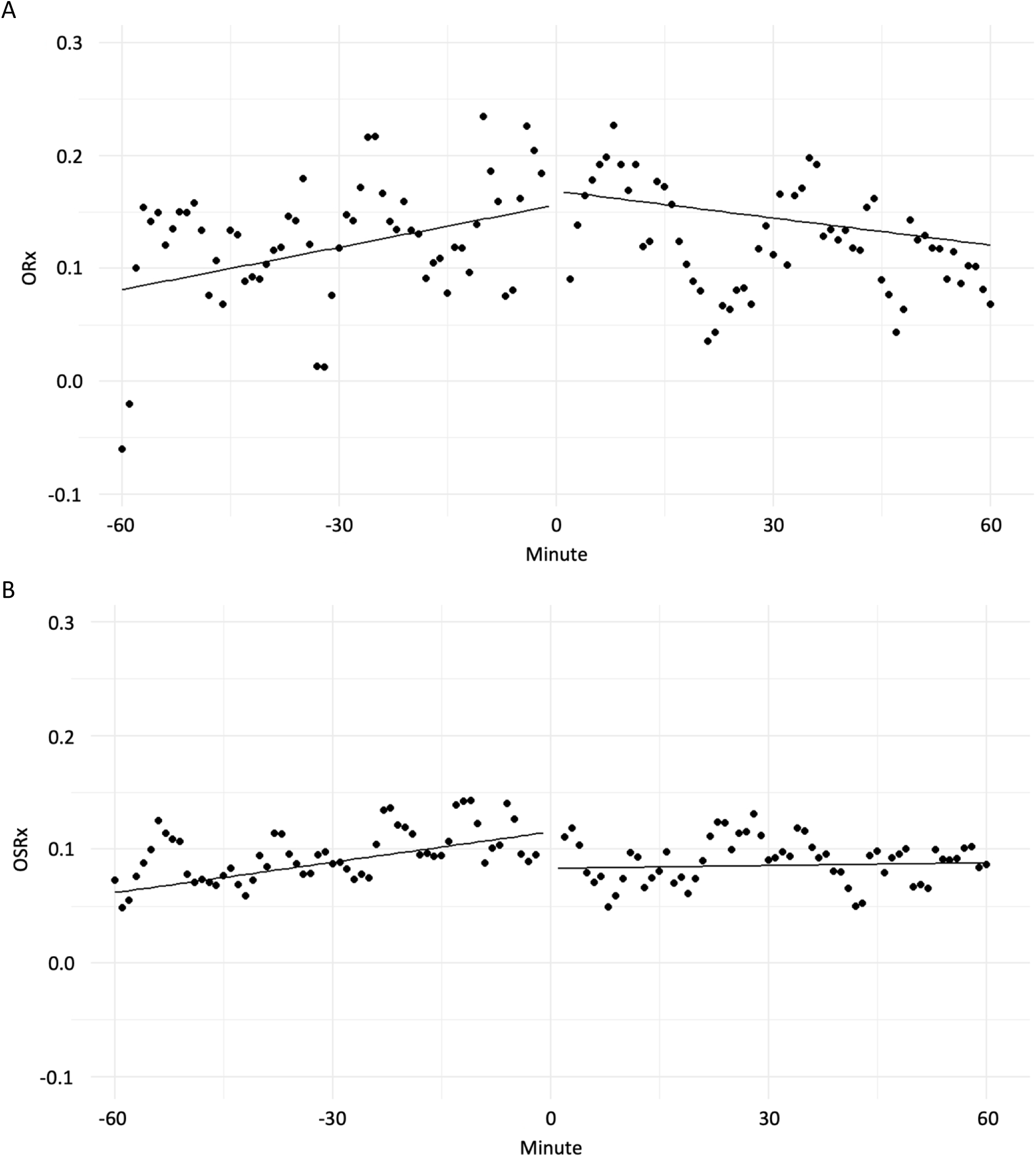
a: Trends in autoregulation index ORx in the 60 minutes pre-SD and 60 minutes post-SD. 4b: Trends in autoregulation index OSRx 60 minutes pre-SD and 60 minutes post-SD. Dots are 1-minute bin average across all SD clusters. Lines are predicted mean values using linear mixed-effects model based on serial data of individual SD clusters (See table 2 for intercept and slope). Individual index values were first transformed using Fisher’s transformation for correlation coefficient.

**Table 2.** Pre- and Post-SD Trend in Autoregulation Based on Linear Mixed Effects Models fit to serial 1-minute bin data up to 60 minutes before and after a SD cluster. Bold/italic values are significant. Grey boxes indicate consistent trends in direction of slope and significance.

| Autoregulation Index | Cluster | Pre-SD |  | Post-Sd |  |
| --- | --- | --- | --- | --- | --- |
|  |  | Slope (/min) | p-value | Slope (/min) | p-value |
| PRx EVD | Single SD | -0.00049 | **0.0349** | 0.00068 | **0.0014** |
|  | Multiple SD | -0.00058 | 0.0556 | 0.00017 | 0.4866 |
|  | Pooled | -0.00052 | **0.0043** | 0.00048 | **0.0025** |
| PRx Parenchymal | Single SD | -0.00040 | 0.1354 | 0.00083 | **0.0021** |
|  | Multiple SD | -0.00152 | **<0.0001** | -0.00026 | 0.4324 |
|  | Pooled | -0.00082 | **0.0001** | 0.00041 | **0.0510** |
| ORx | Single SD | 0.00124 | **0.0070** | -0.00009 | 0.8376 |
|  | Multiple SD | 0.00133 | **0.0463** | -0.00217 | **0.0007** |
|  | Pooled | 0.00125 | **0.0010** | -0.00080 | **0.0288** |
| OSRx (average R/L) | Single SD | 0.00089 | **0.0017** | -0.00003 | 0.9265 |
|  | Multiple SD | 0.00088 | **0.0221** | 0.00029 | 0.4199 |
|  | Pooled | 0.00089 | **0.0001** | 0.00008 | 0.7096 |
| CBFRx | Single SD | 0.00049 | 0.4094 | 0.00017 | 0.7835 |
|  | Multiple SD | -0.00195 | **0.0249** | -0.00202 | **0.0062** |
|  | Pooled | -0.00036 | 0.4683 | -0.00058 | 0.2271 |

## Discussion

The importance of autoregulation in the management of aSAH has been previously explored as a strategy to develop individualized management strategies for patients at risk of ischemia related to DCI^33,34^. Current AHA guidelines for the management of aSAH^35^ suggest that after ensuring appropriate euvolemia, permissive autoregulation strategies are reasonable, however, further validation of algorithms and real time application are needed. In the absence of such individualized approaches, blood pressure augmentation in response to neurologic changes related to DCI may be considered^35^. The use of autoregulation-derived approaches potentially offers the opportunity to better refine such augmentation to the physiology of a given patient at a given stage after injury, though evidence for such strategies remains limited^36^. This is likely in part due to multiple different, non-interchangeable methods to assess cerebral autoregulation and a lack of clear understanding of the exact physiology that is being measured by these indices^37^. The current study therefore fills several important missing links in the literature. First, we have compared several measurement approaches side by side in a relatively large cohort of patients and demonstrated consistent association with outcomes with ORx and OSRx, consistent with other reports where the PRx was less strongly associated with outcomes in aSAH^6,38^. Second, our data provides an important mechanistic link to outcomes based on the relationship of impaired autoregulation to SD.

These data serve as a validation of the hypotheses developed in a smaller cohort of patients who were only assessed if each subject had all the autoregulation monitoring approaches (ICP, PBtO2, CBF, and NIRS) including SD monitoring^77^. In that study, we found that ORx, OSRx and PRx were the most reliable indices in identifying the risk of worse outcomes in patients with aSAH. Only one of the two PRx measurements (parenchymal ICP) was associated with clinical outcomes. This PRx measurement was also inconsistently associated with SD occurrence. In the current study, PRx was not associated with outcomes and there was a possible weak association with SD, though was not consistent when considering single versus clustered SD. On the other hand, both ORx and OSRx were associated with clinical outcomes and were found to increase prior to SD regardless of whether considering single or clustered events. A strength therefore of the current analysis is that this larger study replicates and strengthens the results of the exploratory study.

Based on these data, we agree with current evolving management strategies that target optimal MAP or cerebral perfusion pressure in patients with aSAH^33,39^ and also agree with the importance of tissue oxygenation as these studies have emphasized. The use of the ORx or OSRx as the input function for calculation of CPPopt or MAPopt may be a more effective strategy than using PRx and PbtO2 or rSO2 as separate measures to balance. Certainly, further practical studies are needed to determine the feasibility of incorporating such approaches at the bedside, however our work demonstrating this potential application of either invasive or non-invasive approaches may facilitate use in more patients.

The second important link provided by our current analysis is in refining the relationship between autoregulation and SD. Specifically, SD triggered by decreasing CBF and impaired autoregulation may be one of the central mechanisms of secondary ischemia in aSAH. In a recent multicenter study from Europe, the peak total spreading depolarization-induced depression duration of a recording day (PTDDD) was found to predict ischemia and infarction with 60 and 180-minute duration and concluded SD to be an independent mechanistic biomarker for DCI and delayed infarction in aSAH^13^. This is consistent with previous literature linking SD and spreading ischemia to worse outcomes in aSAH^13,23^. In the current study, we once again identified a triphasic probability curve of SD which has now been demonstrated in patients with traumatic brain injury^40^ and ischemic stroke^26^. Overall, it appears that there in increased risk of SD (lower limit of autoregulation) as high as a MAP of 90-100mmHg in this population, however we do not propose that this be used as in indiscriminate target. Instead, the use of the combination of autoregulation and SD monitoring could play an important role in better understanding the risk that a given patient may be at in terms of ischemia. For example, the occurrence of SD may be an indicator of metabolically compromised tissue (at risk of DCI) and progressively worsening duration of the ECoG depression could indicate the need to further optimize physiologic targets to avoid ischemia using MAPopt or other approaches^16^.

Previous data on the relationship between SD and autoregulation was summarized in a recent comprehensive review^16^. In addition to the associations of ORx and OSRx that we previously reported^7^, another group found associations between SD and impaired autoregulation as measured by the PRx^10^, hypothesizing that SD may in fact be the source of impaired autoregulation. Interestingly, these findings were different depending on whether a parenchymal or ventricular source of ICP monitoring was used. In our data, PRx values from both the parenchymal and ventricular source demonstrated an inconsistent relationship with SD. Both tended to decrease prior to SD and generally tended to increase after SD with significant positive slopes for both sources. These PRx trends indicate variability in relationship to SD however could be consistent with the previous associations of SD leading to worsening PRx^10^. Since PRx was not consistently associated with outcome, it seems that the association between ORx and OSRx with SD may be more clinically relevant.

The relationship between SD and autoregulation may also be more complex, as locally impaired autoregulation at the time of SD has also been observed^11,41^. Therefore, SD may both be triggered by globally impaired autoregulation and may further worsen autoregulation in the region of SD as one mechanism of ongoing tissue metabolic stress. This potentially cyclical relationship is therefore one explanation for these seemingly contradictory findings.

### Limitations

While there are notable strengths to this study including high-resolution recordings over a long period of time in a large, relatively homogeneous cohort of aSAH patients, there are clearly some limitations. Not all subjects had all parameters measured due to practice and technology changes over the course of monitoring. A relatively small number of only surgical patients had SD monitoring, so it is unknown if our observations related to SD apply to non-surgical patients as well, however it seems plausible that similar pathophysiology is at play. In addition, there is a risk that the continuous indices do not reflect the tissue most at risk for SD since the monitors used to generate these indices are typically placed contralateral to the surgical site and therefore the SD monitoring electrode site. Finally, since this was primarily a cohort of more severely injured subjects requiring multimodality monitoring, the applicability to lower-grade patients is unknown.

## Conclusion

Impairment in cerebral autoregulation in aSAH is most associated with worse clinical outcomes and occurrence of SD when using ORx and OSRx. Impaired autoregulation appears to precede SD occurrence rather than be a result of SD occurrence, though this could be a cyclical relationship. Targeting the optimal MAP or cerebral perfusion pressure in patients with aSAH should use ORx and/or OSRx as the input function rather than intracranial pressure. The combined use of continuous autoregulation monitoring to determine the optimum blood pressure target with monitoring of SD as a mechanism of ongoing metabolic stress and risk of ischemia may be a rational strategy to improve outcomes in patients with aSAH.

## Data Availability

Data are available with appropriate data use agreements and institutional approvals

## Acknowledgements

none

